# Parallel detection of multi-contrast MRI and Deuterium Metabolic Imaging (DMI) for time-efficient characterization of neurological diseases

**DOI:** 10.1101/2023.10.02.23296408

**Authors:** Yanning Liu, Henk M. De Feyter, Zachary A. Corbin, Robert K. Fulbright, Scott McIntyre, Terence W. Nixon, Robin A. de Graaf

**Author notes:** Address of correspondence: Yanning Liu, Magnetic Resonance Research Center, Department of Radiology and Biomedical Imaging, Yale University School of Medicine, 300 Cedar Street, N147, New Haven, CT 06520-8043, USA.

## Abstract

Deuterium Metabolic Imaging (DMI) is a novel method that can complement traditional anatomical magnetic resonance imaging (MRI) of the brain. DMI relies on the MR detection of metabolites that become labeled with deuterium (^2^H) after administration of a deuterated substrate and can provide images with highly specific metabolic information. However, clinical adoption of DMI is complicated by its relatively long scan time. Here, we demonstrate a strategy to interleave DMI data acquisition with MRI that results in a comprehensive neuro-imaging protocol without adding scan time. The interleaved MRI-DMI routine includes four essential clinical MRI scan types, namely T_1_-weighted MP-RAGE, FLAIR, T_2_-weighted Imaging (T_2_W) and susceptibility weighted imaging (SWI), interwoven with DMI data acquisition. Phantom and in vivo human brain data show that MR image quality, DMI sensitivity, as well as information content are preserved in the MRI-DMI acquisition method. The interleaved MRI-DMI technology provides full flexibility to upgrade traditional MRI protocols with DMI, adding unique metabolic information to existing types of anatomical image contrast, without extra scan time.

## Main

Magnetic Resonance Imaging (MRI) is the most widely used imaging method in neurology and neurosurgery^1,2^. This non-invasive and non-radioactive technique leverages signal from the nuclear spin of water protons to provide high-resolution and high-contrast images. Yet, conventional MRI scans depict primarily anatomical and physiologic information and can have limited specificity when imaging neurological diseases^3-5^. In brain tumors, for example, determining tumor grade, and distinguishing tumor recurrence from treatment-induced changes using MRI is a longstanding challenge. These limitations in characterizing neurological lesions can significantly impact clinical decision-making; therefore, it is essential to explore alternative imaging approaches to overcome these issues.

To enhance clinical assessment of central nervous system (CNS) diseases, it is imperative to harness information beyond abnormal anatomy and physiology. As our understanding of brain diseases deepens, it has become clear that alterations in metabolism is a key feature of numerous neurological disorders.^6-10^ For brain tumors, cancer-associated metabolic reprogramming, including altered glucose metabolism, is increasingly recognized as a hallmark of the disease.^9^ Consequently, the availability of a medical imaging method that provides insights into metabolism would greatly support clinical decision-making in the context of CNS diseases.^3,4,11^

The use of isotopically labeled substrates offers a method to image active metabolism in vivo. Positron emission tomography (PET) of the glucose analog deoxyglucose (DG) labeled with the radiotracer ^18^F is the most prominent metabolic imaging approach in the clinic, particularly in the setting of cancer. An elevated ^18^FDG uptake indicates an increasing demand of glucose, which is often correlated with a specific type of glucose metabolism observed in aggressive tumors (the Warburg effect). As a result, ^18^FDG-PET is successfully used to characterize various cancers outside of the brain. Nevertheless, in patients with brain tumors, the high baseline glucose uptake in healthy brain can lead to low tumor-to-brain image contrast, often resulting in inconclusive ^18^FDG-PET scans^4,12,13^. In addition, the exposure to ionizing radiation during ^18^FDG-PET complicates repeated scanning as well as the use in pediatric patients. Deuterium Metabolic Imaging (DMI)^14^ is a recently developed method to map active metabolism in vivo. It offers an alternative approach to image glucose metabolism in neurological diseases because it uses non-radioactive, deuterium labelled substrates and provides high contrast in the brain. After oral administration of [6,6’-^2^H_2_] glucose solution (by drinking), DMI can quantitatively detect the labeled glucose (Glc) as well as its downstream metabolites, ^2^H-labeled lactate (Lac) and glutamate + glutamine (Glx), in vivo. The resulting maps of the individual metabolites, and combinations thereof, present distinct types of image contrast that highlight differences in glucose metabolic pathways used by normal brain and lesions caused by CNS diseases^14,15^. The potential for high image contrast with normal brain, the use of a stable, non-radioactive isotope and the relative simplicity of the technique make DMI attractive for clinical implementation.

A single complete DMI measurement typically requires approximately 30 minutes to achieve sufficient sensitivity in vivo. Simply adding a DMI scan to existing neuro-MRI protocols will significantly lengthen the total scan time, compromising patient adherence and scanner throughput. A time-efficient strategy that combines DMI with traditional MRI will facilitate the introduction of DMI to the clinics. Interleaved multinuclear acquisition^16-18^ is a technique that successively measures magnetic resonance signals from more than one nucleus by alternating the Larmor frequency of detection within a short time period, typically less than one pulse sequence repetition time (TR). This approach offers an opportunity to incorporate DMI pulse-acquisitions into MRI schemes, thereby acquiring both ^1^H and ^2^H signal, whose resonance frequencies are 6.5 times apart from each other. Additionally, interleaved acquisition provides a high level of flexibility in designing RF pulse sequences and can be implemented on MRI scanners with minimal hardware modifications^17,19^.

Earlier we have described the hardware modifications and demonstrated the proof-of-principle of parallel MRI-DMI using the interleaved ^1^H^2^H methodology, with a single type of MRI pulse sequence^20^. However, clinical neuro-radiology MRI protocols include multiple types of MRI scans to generate different tissue contrasts. In this work, we set out to build a comprehensive MRI-DMI protocol that encompasses a series of routine clinical MRI scans, including T_1_-, T_2_-weighted imaging and susceptibility weighted imaging (SWI), while satisfying the following goals: (1) the MRI contrasts and sensitivities remain unchanged. (2) the total duration of the scan sessions cannot increase by more than 10% (as compared to the standard, mononuclear MRI), and (3) DMI sensitivity and resolution are preserved. The resulting routine, consisting of five parallel MRI-DMI methods, generates four MRI contrast types in parallel to DMI in 42 minutes. (**Fig. 1a**) We first validated the method in phantoms, followed by detecting natural abundant HDO in healthy volunteers (N=3), and ^2^H-labeled glucose metabolites after oral intake of [6,6’]-glucose (N=2). Finally, the MRI-DMI method was used to acquire high quality MRI and DMI in a patient with a high-grade brain tumor.

**Figure 1.**
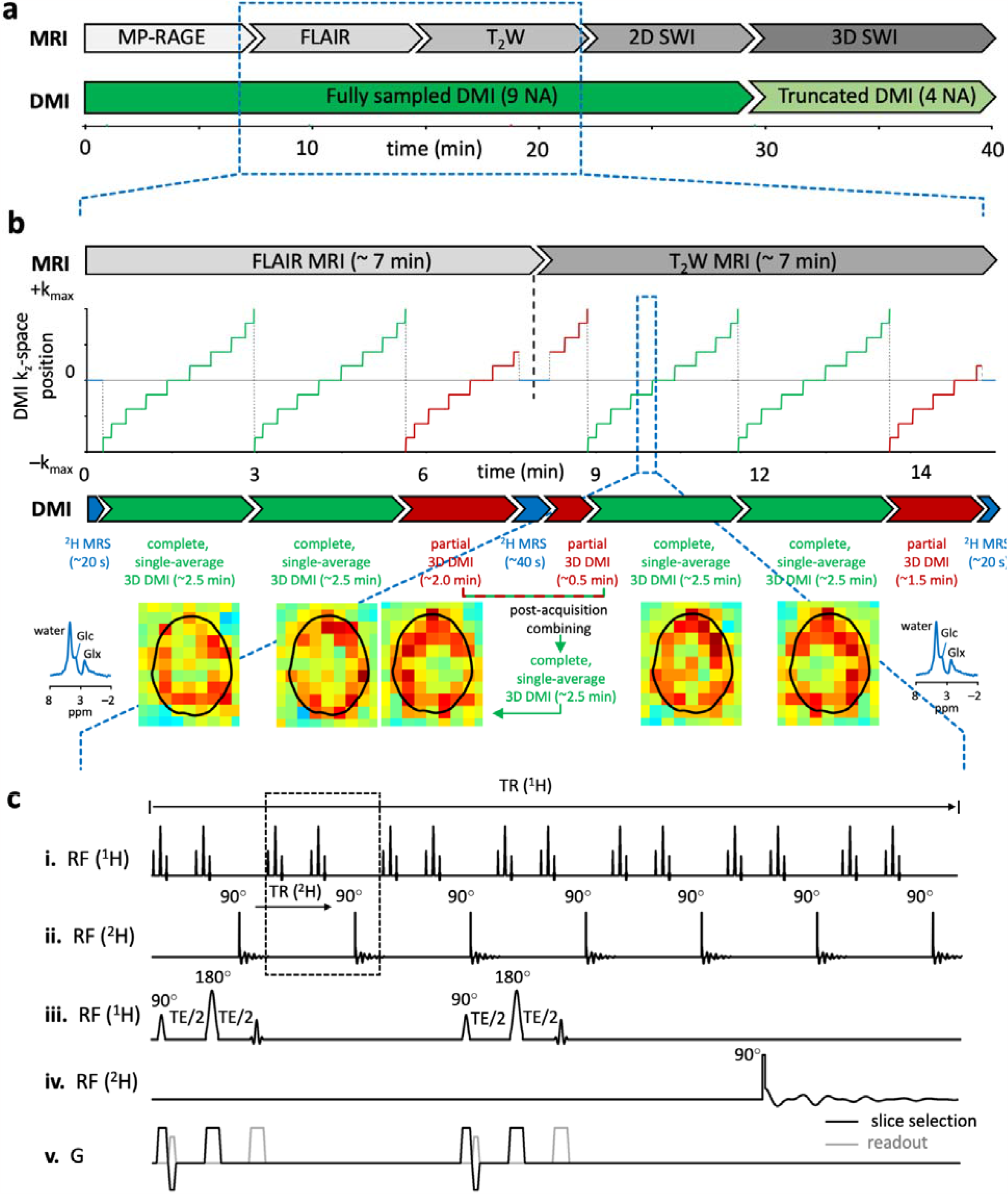
Schematic of the parallel, multi-contrast MRI and DMI brain imaging protocol. (**a**)The overall imaging routine includes five parallel MRI-DMI scans, namely MP-RAGE-,FLAIR-, T_2_W-, 2D and 3D SWI-DMI. This 42-minute scanning procedure enables nine averages of fully sampled DMI and four averages of truncated DMI without increasing the standard MRI-only scan time. **(b)** The parallel acquisition of ^1^H MRI signal is essentially identical to that during a standard, proton-only method. For the example shown, FLAIR and T_2_W MRI datasets are both acquired in ∼7 min. In parallel to the MRI data, a single-average 3D DMI dataset can be acquired in ∼2.5 min. Two complete 3D DMI datasets (green periods indicated in the k_z_-space position graph. The corresponding k_x_ and k_y_ positions are omitted for visual clarity) can be acquired during the FLAIR method. The third average is incomplete (red period) as the FLAIR MRI acquisition ends earlier. The missing k-space encodings from the third average can be obtained at the beginning of the following scan, T_2_W-DMI. The two partial 3D DMI datasets can be combined post-acquisition to give a complete 3D DMI dataset. Global ^2^H MR spectra (blue periods, no phase-encoding) are acquired for a short period at the start and end of each MRI method to (1) monitor metabolic stability, frequency drifts and overall spectral quality and (2) reestablish a steady-state for the ^2^H magnetization at the start of a new MRI sequence. **(c)** Pulse sequence diagram of parallel T_2_W MRI-DMI. ^2^H pulse acquisitions are interleaved into the intrinsic time delays of ^1^H MRI sequences. **(i/iii**) 14 ^1^H MRI slices are excited and refocused (TR/TE = 2200/40 ms) to form spin echoes. A temporally non-uniform ^1^H slice excitation is used to accommodate DMI insertion. **(ii/iv)** ^2^H pulse acquisitions are placed every 2 excitations to maintain a DMI TR of 314 ms. **(v)** Shows the gradients used during acquisition.

## Results

Five MRI methods, providing unique and clinically relevant image contrasts, were successfully interleaved with parallel DMI acquisitions. Each parallel method was designed to be executed individually with its independent parameter control interface. **Fig. 1a** shows the overall timing of the five-sequence protocol over a 42 min period, together with the interleaved DMI acquisitions. Even though the 2D and 3D SWI methods generate similar image contrast, they were both included to highlight the sequence design considerations between 2D multi-slice and 3D interleaved MRI-DMI methods. The time duration of an MRI-DMI scan is governed by the MRI acquisition scheme, giving rise to partial, incomplete DMI datasets towards the end of each method. **Fig. 1b** demonstrates how partial DMI acquisitions started during one parallel scan can be completed during a following MRI-DMI scan, allowing for near-continuous DMI signal averaging during the entire imaging protocol. **Fig. 1c** provides the scheme to interleave DMI with the multi-slice, T_2_-weighted MRI, whereby temporally non-uniform ^1^H slice acquisitions generate sufficient interslice delay to allow a ^2^H pulse-acquire acquisition after every two ^1^H slices to provide a constant ^2^H repetition time. **Fig. S1** shows detailed pulse sequence diagrams for parallel MP-RAGE-DMI (**Fig. S1a**), 2D multi-slice SWI-DMI (**Fig. S1b**) and 3D SWI-DMI (**Fig. S1c**). Each MRI sequence requires specific considerations and modifications to allow the placement of ^2^H excitation and acquisition blocks at a constant TR, which are described in depth in the Methods.

The collection of parallel MRI-DMI methods shown in **Fig. 1a** was rigorously tested on phantoms in vitro, whereby parallel and sequential (i.e. standard) acquisitions were compared for SNR of ^1^H and ^2^H signal, and for spectral resolution of ^2^H. The SNR of parallel MRI acquisition was 100 ± 5 % of the standard MRI acquisitions, as averaged over all five MRI methods. ^1^H SNR comparisons for individual MRIs are summarized in **Fig. S2e**. The parallel and standard MRIs showed high correlation and agreement, with random noise being the main source of variability between the measurements (**Fig. S3**).

Interleaved global ^2^H MRS signals were compared with its standard ^2^H-only reference (**Fig. S2f**). The SNR of ^2^H signal from five individual parallel scans showed no significant difference from their control. All parallel ^2^H MRS data was then summed following the inter-sequence “stitching” algorithm as described in the Methods. The SNR of combined global ^2^H signal was comparable with the standard ^2^H-only control, ensuring ^2^H signal from different parallel scans can be effectively summed without losing sensitivity. The SNR of DMI acquired using the 5-sequence routine was 98 ± 4 % and 99 ± 4 % for water and DMSO, respectively, of standard (mono-nuclear) DMI (Fig. **S2g)**. The ^2^H spectral linewidths (LW) of water and DMSO were comparable between standard and parallel acquisitions (98 ± 2 % and 96 ± 3 % for water and DMSO, respectively), indicating a negligible effect of MRI-related eddy currents on the performance of DMI (**Fig. S2h)**. The marginally higher linewidth in standard DMI could be attributed to the B_0_ frequency drift over the measurement time, which was corrected during the inter-sequence “stitching” of parallel DMI, as described in the methods.

The performance of parallel MRI-DMI was evaluated on healthy volunteers at natural abundance (N = 3) and 60-120 min following oral [6,6’-^2^H_2_]_-_glucose administration (N = 2). Parallel (interleaved) and standard (direct, mono-nuclear) MRIs (**Fig. 2a**) all displayed expected image contrasts and excellent image quality. The contrasts of 2D and 3D SWI were different, as expected, primarily due to increased T_1_ weighting during the 3D acquisition. The visual correspondence between the direct and interleaved MRIs was further corroborated by a quantitative comparison based on image SNR and contrast (**Fig. 2c**) across segmented gray matter (GM), white matter (WM) and cerebrospinal fluid (CSF) regions (**Fig. 2b**). Good correlation and agreement were observed between image pairs, despite a slightly higher variability as compared to the experiments in phantoms, due to subtle motions (i.e. movements, breathing, CSF pulsation etc.) inherent to in vivo scans. (**Fig. S4**)

**Figure 2.**
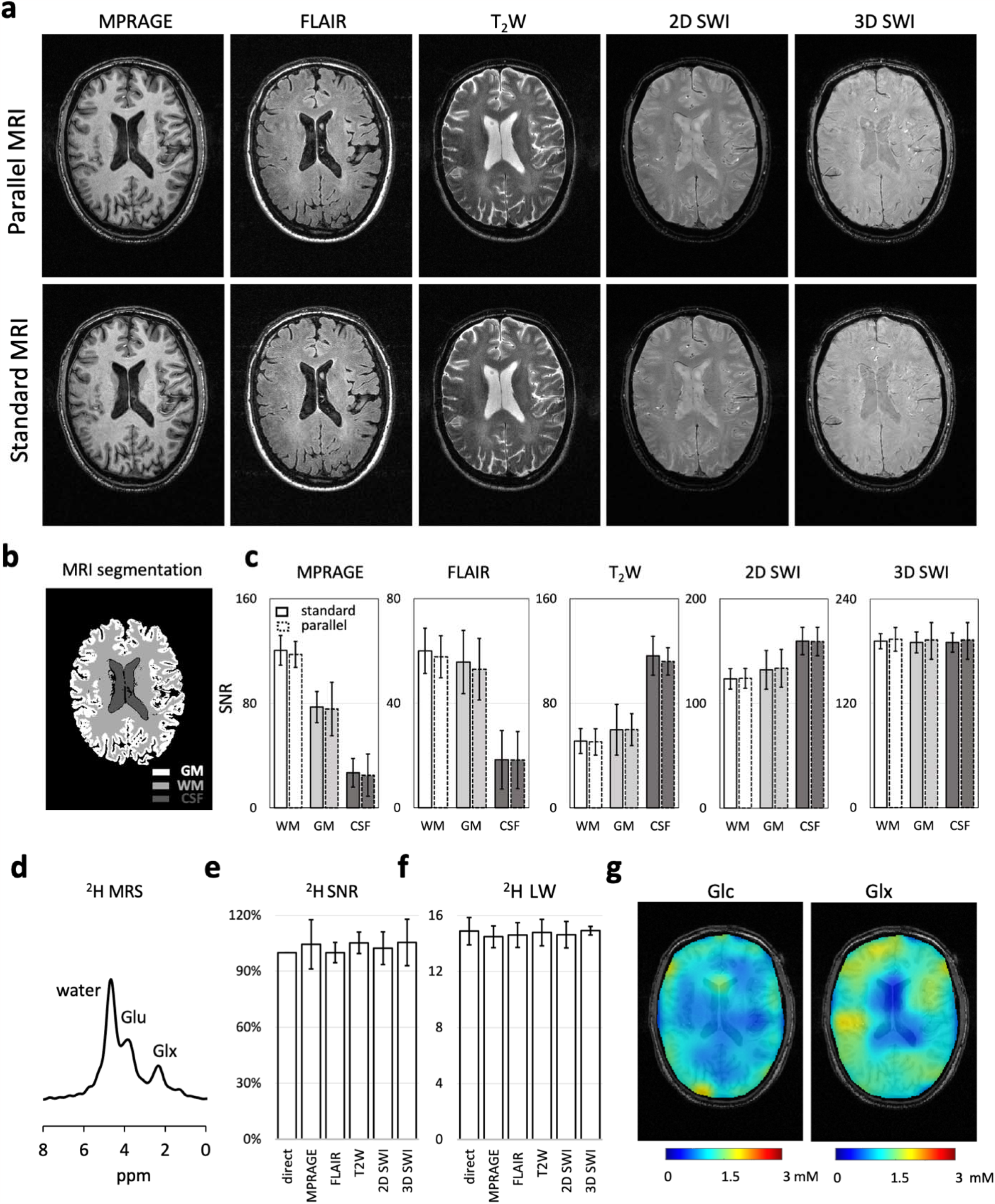
Parallel MRI-DMI on a healthy brain. **(a)** Parallel and standard MP-RAGE, FLAIR, T_2_W, 2D SWI and 3D SWI MRIs acquired of a healthy brain. (**b**) The brain was segmented into white matter (WM), gray matter (GM) and cerebrospinal fluid (CFS) based on the MP-RAGE image. **(c)** The parallel versus standard MRI SNR comparisons in WM, GM and CSF are shown on the displayed brain slice. The error bars represent the standard deviation of image pixels(/voxels) in each segmented brain area. **(d)** The global ^2^H MR spectrum, acquired in parallel to the interleaved MRIs in **(a)**, is shown. **(e-f)** The SNR and LW (of water peak) in the global ^2^H MR spectra are compared between the parallel and standard acquisitions (N=3). **(g-h)** DMI concentration maps of ^2^H-labeled Glc and Glx normalized to water, based on spectral fitting, assuming the concentration of deuterium in water remains constant across voxels (10 mM). One-way or two-way ANOVA were used for statistical analysis. No significant difference (P<0.05) was found between standard and parallel measurements in **(c), (e)** and **(f)**.

Following basic processing, DMI can be transformed into 3D metabolic maps of the ^2^H-labeled substrate, [6,6’-^2^H_2_]-glucose and the metabolic product, [4-^2^H]-glutamate and [4-^2^H]-glutamine (**Fig. 2g**). In agreement with previous reports^14^, the Glc and Glx maps are relatively uniform, with a noticeable lack of Glx in the ventricles where glucose metabolism is expected. In healthy brain, the metabolic product [3-^2^H]-lactate is typically near the noise floor (not shown). The ^2^H water spectral linewidths (**Fig. 2f**) and SNR (**Fig. 2e**) are constant across different MRI methods, in close agreement with the phantom results (**Fig. S2**), although with slightly higher variation as a expected.

**Fig. 3** shows results from the parallel MRI-DMI brain imaging protocol as applied in a patient with an IDH-mutant astrocytoma, WHO CNS Grade 4, brain tumor 80-120 minutes after ingestion of [6,6’-^2^H_2_]-glucose. There is excellent contrast between tumor and normal brain on FLAIR and T_2_-weighted MRI (**Fig. 3a**), a result of infiltrating tumor and tumor-associated edema. The 2D and 3D SWI sequences also demonstrate good contrast between the tumor and normal brain, indicating that the tumor has areas of increased susceptibility. . While the T_1_-weighted MP-RAGE image shows excellent GM/WM contrast, tumor and normal brain contrast was less pronounced, which is typical in the absence of T_1_-enhancing contrast agents. DMI-extracted ^2^H MR spectra (**Fig. 3b**) and DMI-derived maps of Glc, Glx and Lac (**Fig. 3c**), acquired in parallel with the multi-contrast MRIs, display significant metabolic contrast. A drastic decrease of Glx and an appearance of Lac were observed in the tumor lesion as a result of cancer-associated metabolic reprogramming. The Lac/(Lac+Glx) ratio illustrate the extent of the Warburg effect, thereby exquisitely differentiating tumor from healthy brain tissues.

**Figure 3.**
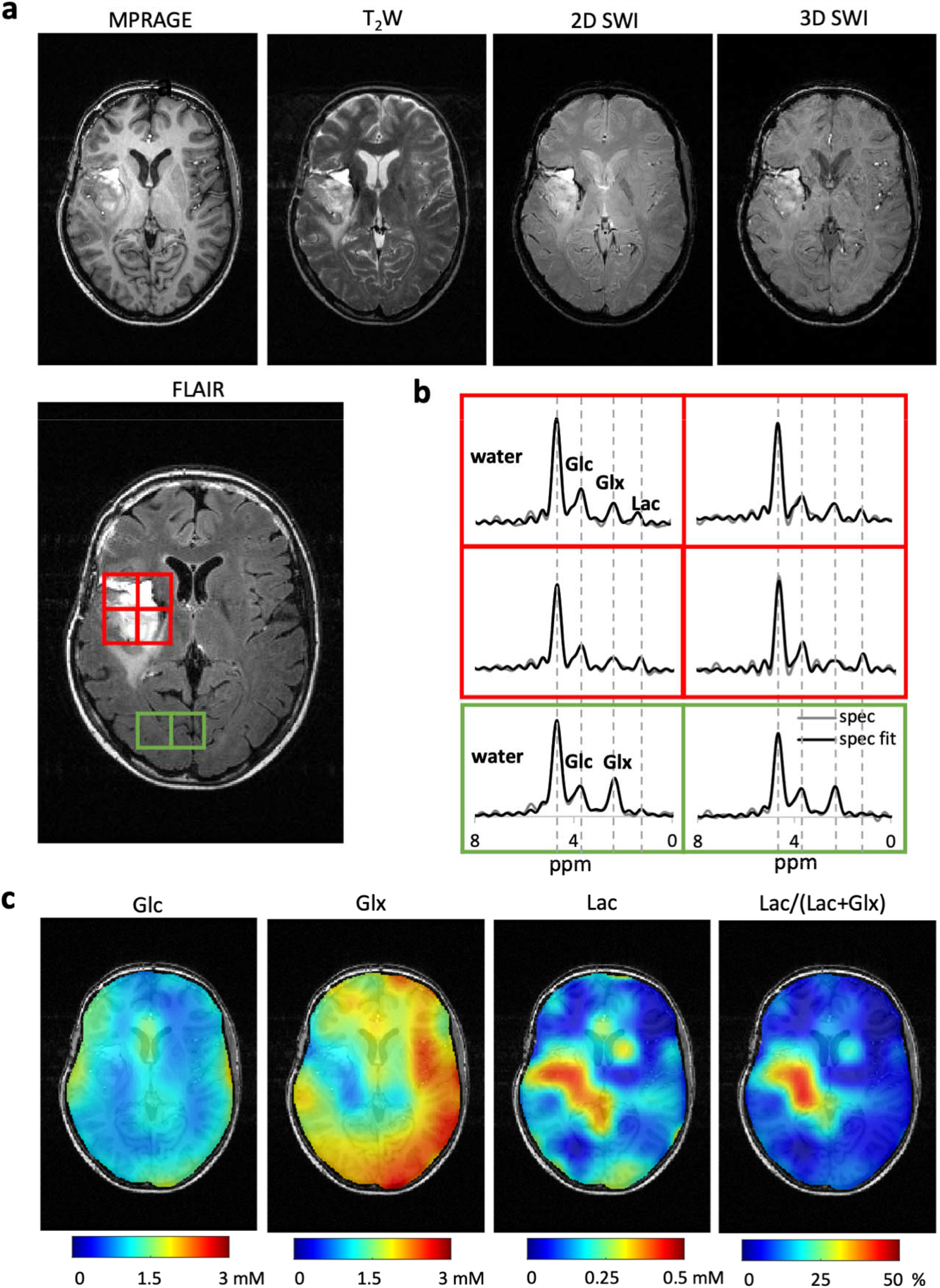
Parallel MRI and DMI on a brain tumor patient. **(a)**. MRIs acquired using the MRI-DMI protocol demonstrates the tumor area in the brain. Representative leisional and healthy brain tissue regions are highlighted on the FLAIR MRI in red and green boxes respectively, with their DMI spectra displayed in **(b). (c)** Metabolic maps representing the concentration of Glucose (Glc), Glutamate +Glutamine (Glx) and lactate (Lac) obtained using spectral fitting. The Lac/(Lac+Glx) ratio is used to illustrate the extend of the difference in glucose metabolism, differentiating tumor from healthy brain tissues.

Deuterium-based spectra and maps data presented in **Fig. 3** were based on nine DMI datasets with full-length FIDs (T_acq_ = 80.5 ms) acquired during MP-RAGE, FLAIR, T_2_W and 2D SWI and four DMI datasets with truncated FIDs (T_acq_ = 25 ms) acquired during 3D SWI. The ability to separate the ^2^H-labeled signals of water, Glc and Glx strongly depends on the FID length and simulations show that the Cramer-Rao lower bounds (CRLBs) for all signals rapidly increase for acquisition times less than 30-40 ms. (**Fig. S5**) While the truncated DMI data by itself is insufficient to reliably separate the spectroscopic ^2^H signals, adding these data to full-length DMI data provides an average of 15% of SNR boost on in vivo data (**Fig. S6c**). This represents ∼95% of the theoretical maximum (if all ^2^H FID were acquired in full length), consistent with Monte Carlo simulation results (**Fig. S4a-b**).

## Discussion

Energy metabolism is essential to brain function and its disruption is closely associated with various CNS diseases. A time-efficient and robust method to supplement clinical brain MRIs with regional information of active metabolism could significantly improve the clinical evaluations of these diseases. To address this need, we developed a parallel multi-contrast MRI and DMI protocol that allows the acquisition of clinical brain MRI scans, in addition to metabolic imaging via DMI, using interleaved ^1^H/^2^H technology. The routine acquired four types of clinical MRI contrasts, namely MP-RAGE, FLAIR, T_2_W and SWI (2D and 3D), and a complete set of DMI data with high SNR in 42 minutes, without lengthening the original MRI scan time. The MRI sequences were selected to match the clinically relevant scans for the diagnosis and management of brain tumors and neurological diseases.^1,21^ DWI, a prevalently used MRI scan, was not implemented here due to the limited slew rate and amplitude of the gradient coils on our research MRI scanner. However, the T_2_W method is essentially the equivalent of a low-b-value DWI method and can be readily converted into DWI with a wide range of choices on echo time (TE) and b-value. The parallel MRI-DMI protocol was validated on MRI sensitivity and contrast and DMI sensitivity and resolution, on phantoms in vitro and human brains in vivo. Following validation, the routine was successfully applied to human brains following the administration of deuterated glucose, including in a patient with an IDH-mutant astrocytoma, WHO CNS Grade 4, brain tumor. In all cases, the parallel MRI-DMI acquisitions showed high-quality, artifact-free MRIs and DMI data with the typical spectroscopic fingerprint of water, Glc and Glx. In the brain tumor patient, the pathological region was not only identified with expected MRI contrast, but also clearly highlighted with a metabolic alteration, showing increased Lac and decreased Glx levels.

To harness the maximal DMI sensitivity, we employed a continuous DMI phase-encoding scheme between consecutive scans, facilitating the stitching of partial DMI datasets. Non-phase-encoded DMI acquisitions at the beginning and the end of each MRI method ensured phase continuity, making this approach generally applicable to any combination of MRI sequences. For the presented five MRI-DMI protocols, the overall DMI acquisition duty cycle (TR = 314 ms) was 90.6% of the theoretical maximum. MRI methods without sufficiently long delays to allow complete DMI sampling (e.g., 3D SWI) were modified to enable the acquisition of truncated DMI signals. While the truncated signals by themselves provided insufficient data quality, as quantified by the high CRLBs (**Fig. S5**), the combined fitting of truncated and full-length DMI data provided a significant boost in DMI sensitivity over simply omitting the truncated data (**Fig. S6)**.

Any MR pulse sequence is limited by interdependent parameter settings (e.g., TE < TR). For example, in a multi-slice, T_2_-weighted MRI-only method the maximum number of slices, NS, is limited by TE and TR. For a given TR, NS can increase by decreasing TE, possibly down to the minimum TE given by RF pulse lengths, gradient crusher durations and acquisition lengths. This common correlation between multiple parameters is governed by calculations running in the background that present the MR operator with parameter ranges aimed at maintaining non-negative delays throughout the method. Parallel MRI-DMI methods will have a similar interplay between parameters, whereby ^1^H and ^2^H-specific conditions must be attained simultaneously. Many parameters, such as MRI slice thickness, in-plane geometry, or image orientation, will have identical ranges to MRI-only methods, whereas other parameters, such as NS, can be limited to a smaller range. The slightly reduced parameter space is, however, offset by the simultaneous acquisition of DMI-based metabolic information. For both MRI-only and parallel MRI-DMI methods, the MR operator sees a similar user interface which only presents valid parameter choices. As is common practice for MRI-only methods, different MRI contrast mechanisms (e.g., T_1_, T_2_, SWI) can be executed with separate parallel MRI-DMI methods, each with their own set of unique parameter conditions (**Figs. 1c and S1**).

The single-channel Transverse ElectroMagnetic (TEM) ^1^H volume coil used in the presented studies, prevented the implementation of commonly used acceleration methods such as SENSE^22^ or GRAPPA^22^. However, the developed methods are fully compatible with the reduced encoding associated with parallel imaging. When the DMI sensitivity is sufficiently high (e.g., at 7T^23^ or beyond^24^) the acceleration can be used to achieve a shorter overall scan duration. When the full 30-40 min duration is needed for extended DMI signal averaging (e.g., at 3T or 4T), the accelerated MRI acquisition can be complemented with additional MRI contrasts, like CEST or DWI, or with other spectroscopic measurements, such as ^1^H MRSI^25^.

The multitude of potential MRI contrasts has led to a wide range of MRI acquisition methods. While it has been demonstrated here that many different MRI sequence types can be modified to allow parallel acquisition of DMI, it is challenging to formulate a universal approach for incorporating DMI elements into MRI methods. Nevertheless, there are several guidelines that can be followed to convert MRI-only methods in parallel MRI-DMI sequences. Strategies to achieve the overall goal of unchanged MRI contrast and a constant DMI TR, include (A) ^2^H pulse acquire windows placed within the inherent time delays of MRI (e.g. DMI during FLAIR),(B) during periods of dense ^1^H MRI acquisitions, it may be necessary to apply ^2^H RF pulses (without acquisition) to maintain a steady-state for ^2^H longitudinal magnetization (e.g., DMI during MP-RAGE). (C) For multi-slice MRI acquisitions, temporally non-uniform slice excitations may be required to accommodate DMI (e.g., DMI during T_2_W or 2D SWI). (D) To maximize DMI sensitivity, DMI acquisitions may need to be divided across multiple MRI methods, through continuous signal averaging over the entire MRI protocol. (E) In fast MRI sequences, truncated FID can be acquired and combined with fully-sampled DMI obtained during other MRI sequences, contributing to the overall sensitivity (e.g., DMI during 3D SWI).

In addition to the outlined modifications, it is important to realize that unlike nucleus-specific acquisitions, magnetic field gradients affect all spins in the sample simultaneously. Wherever possible, magnetic field gradients required for one nucleus were applied when the magnetization for the other nucleus is along the longitudinal axis. Nevertheless, eddy currents from previously applied gradients can propagate across MRI or DMI time periods and lead to undesirable phase effects. In cases where this effect was observed, magnetic field gradient amplitudes and/or placement were modified to minimize eddy-current-related phase effects.

Multi-nuclear NMR is an excellent strategy to reduce total scan time without sacrificing data quality. While the application of interleaved acquisitions for human clinical research has been a recent development, the prospect to acquire comprehensive information originated from more than one nucleus has generated significant interest.^17,26-28^ One challenge in applying interleaved ^1^H/^2^H technology on the current iteration of modern clinical MRI scanners is that they are optimized to receive signal at one Larmor frequency (i.e. ^1^H) within a given RF pulse sequence. A viable solution, requiring minimal hardware adjustments, involves employing a frequency-conversion device to match the frequency of the nuclei of interest.^19^ interleaved acquisitions in clinical MRIs has also prompted the The growing interest in vendors to incorporate interleaved acquisition options into their systems^29^, reflecting the evolving landscape of MRI technology.

Interleaved ^1^H/^2^H technology provides a unique opportunity to achieve acquisition on both nuclei in parallel. Apart from supplementing clinical MRI to aid the characterization of CNS diseases, the availability of active metabolic information can also facilitate numerous neuroscience research studies. For example, combining DMI with functional MRI will allow researchers to explore the connection between brain function and metabolism. Additionally, DMI can complement ^1^H MRSI by providing a means to compare biochemical data and active metabolism in the brain, enabling more thorough investigations into both normal and pathological brain biochemistry. The demonstration of excellent performance of the interleaved MRI-DMI protocol without a time penalty should accelerate the adoption of DMI for use in clinical settings.

## Methods

### MR system and hardware modifications

A detailed description of the system, including the hardware modifications, has previously been reported.^19^ In short, measurements were obtained on a 4T magnet (Magnex Scientific Ltd.) interfaced to a Bruker Avance III HD spectrometer via ParaVision 6 (Bruker BioSpin, Billerica, MA, USA). RF transmission and signal reception were performed with a quadrature TEM coil for proton and a four-channel trans-receiver array for deuterium. To enable parallel ^1^H and ^2^H signal detection, a customized frequency-conversion unit^19,20^ was used to upconvert the ^2^H signal (26 MHz) to the ^1^H frequency range (170 MHz), after which both ^1^H and the converted ^2^H signals entered the standard ^1^H receive path. Frequency and phase locking of the ^2^H signal were maintained through sequence-specific look-up tables.

### Parallel MRI-DMI sequences

Deuterium windows with complete ^2^H signal acquisition (T_acq_ = 80.5 ms) were interleaved with FLAIR^20^, MP-RAGE **(Fig. S2a)**, T_2_W **(Fig. 1c)** and 2D SWI MRI **(Fig. S2b)** sequences, using the time delays inherent to each MRI method while maintaining a constant DMI TR of 314 ms. A detailed summary of sequence parameters is reported in **Table S1**. The parallel FLAIR-DMI method was previously described^20^ and used without modification. MP-RAGE (TR/TI_eff_ =3140/1240 ms) was collected in 3D as a 256 x 192 x 64 matrix over 256 x 192 x 128 mm^3^, with inversion recovery followed by a collection of 64 rapidly acquired gradient-echoes (TE/ESP=8/20 ms, Flip Angle (FA) =15°). Six DMI pulse-acquire elements (TR=314 ms) were incorporated during inversion recovery and relaxation delays in each ^1^H TR. As the dense ^1^H acquisition period of MP-RAGE did not allow the incorporation of DMI pulse-acquire elements, four equidistant ^2^H excitation RF pulses (TR = 314 ms) were applied to maintain the ^2^H signal steady-state. Multi-slice 2D T_2_W (TR/TE = 2200/40 ms) and 2D SWI (TR/TE = 628/18.5 ms, FA=45°) with flow compensation, were collected as a 256 x 192 matrix over 256 x 192 mm^2^, with 14 slices of 3 mm thickness. Within a given TR, the excitation and acquisition of ^1^H signal for different slices is temporally non-uniform to accommodate the insertion of DMI pulse-acquire elements. For 2D T_2_W and SWI, one ^2^H window is inserted every two spin-echoes (**Fig. 1c**) and every seven gradient-echoes (**Fig. S2b**), respectively. The 3D SWI-DMI (TR/TE=52.4/25ms, FA=25°) was obtained using spoiled gradient echoes, as a 256 x 192 x 64 matrix over 256 x 192 x 128mm^3^ **(Fig. S2c)**. The absence of long delay periods during 3D DWI forced the insertion of truncated ^2^H windows of ∼25 ms (1250 points acquired over 50 kHz). One ^2^H excitation-acquisition was inserted for every six ^1^H excitations to maintain ^2^H TR. For a total SWI acquisition time of 10 minutes and 43 seconds, 4 repeats of truncated DMI were acquired.

DMI was acquired as a 13 x 9 x 11 matrix (YXZ) over 260 × 180 × 220 mm^3^ using spherical k-space encoding, with 491 phase-encoding steps. The parallel MP-RAGE, FLAIR, T_2_W and 2D SWI-DMI phase encoding was continuous between consecutive sequences such that partial DMI datasets from adjacent scans can be adjoined to produce a complete DMI dataset. Non-phase-encoded global ^2^H spectra were acquired in the first and last 55 ^2^H acquisitions of each interleaved sequence to (1) monitor metabolic stability, frequency drifts and overall spectral quality and (2) re-establish a steady-state for the ^2^H magnetization at the start of a new MRI sequence. The entire five-sequence brain imaging protocol (**Fig. 1a**) took 42 minutes in total, giving rise to nine fully sampled DMI repeats and four truncated DMI repeats in addition to the multi-contrast MRIs.

### Phantom Measurements

Phantoms consisting of 1-2 L plastic bottles and spheres and containing ∼0.5% D_2_O and various concentrations of DMSO-D_6_ were employed for all in vitro studies. To allow direct comparison of sensitivity, resolution and artifacts, parallel MRI-DMI acquisition were typically followed by standard, single nucleus MRI and DMI using non-interleaved, mono-nucleus pulse sequences with identical acquisition parameters.

### In vivo Measurements on Healthy Volunteers

All human studies were approved by the Yale University Institutional Review Board. Parallel MRI and DMI were acquired on healthy volunteers without (N=3) or with (N=2) [6,6’-^2^H_2_]-glucose enrichment. In the case of [6,6’-^2^H_2_]-glucose enriched measurements, data were acquired 60-75 min after oral intake of 55 g of [6,6’-^2^H_2_]-glucose dissolved in 250 mL of water, as previously described.^14^ Standard, single nucleus MRI and ^2^H MRS were acquired following the completion of interleaved experiment on the same subjects.

### In vivo Measurements in a Patient with a Brain Tumor

The glucose administration procedure described for healthy volunteers was also used when performing parallel MRI-DMI in a brain tumor patient. To enhance patient comfort and compliance, only the parallel MRI-DMI data were acquired, without a direct comparison to standard acquisition protocols.

### Signal processing

The ^1^H and ^2^H data were individually processed offline in Matlab 2023 (Mathworks, Natick, MA, USA). DMI fitting and data analysis were carried out using an in-house graphical user interface DMIWizard.

### MRI data processing

MRIs were reordered and reconstructed via 2D or 3D Fast Fourier Transform and spatially aligned to the FOV of DMI. Image SNR in vitro was calculated as the absolute-valued pixel intensity divided by the standard deviation of the real component of the background signal. The background signal was chosen from a ROI well outside the phantom, carefully eliminating artifacts from phase-encoding or other origins. Image SNR in vivo was determined for brain white matter (WM), gray matter (GM) and cereberal fluid (CSF), which were determined from the high-contrast MP-RAGE MRI. Image SNR was calculated for all brain regions and for all acquired MRI contrasts.

### DMI data processing

Phase and frequency differences of DMI data acquired between different MRI sequences were corrected using the global, non-phase-encoded ^2^H MR signals obtained at the start and end of each sequence. The full-length ^2^H data obtained in the first four parallel scans (**Fig. 2a**) were phase-aligned and drift-corrected using the global ^2^H MRS obtained during the first and last 55 FIDs in each sequence, before being combined and reconstructed into a DMI dataset (NA = 9). The truncated DMI averages (NA = 4) were summed and reconstructed independently of the fully sampled DMI. The fully-sampled and truncated DMI were then adjoined, fitted using a linear combination of model spectra (LCM). The fitting was conducted via a nonlinear least-squares minimization weighted by 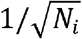, where *N*_*i*_ represents the number of averages of a given data point *i*.^30^ The spatial distribution of Glc (glucose), Glx (glutamate+glutamine) and Lac (lactate) were obtained by analyzing the LCM spectral fitting results. The Glc and Glx concentration maps were calculated assuming a spatially homogenous deuterium concentration in water (10 mM), corrected for T_1_ and B_1_ effects and ^2^H label loss due to exchange^31^.

To present the resulting DMI on the spectral domain, the averaged FID, *s*_*avg*_ (*t*), was calculated by averaging both the truncated and fully-sampled DMI:

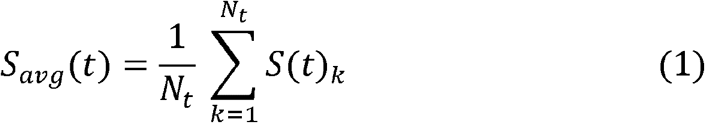

where *N*_*t*_ equals to the total number of averages acquired at time point *t*.

Following Fourier transform, the signal-to-noise ratio (SNR) was calculated as 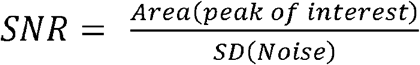, where the area under the peak is extracted from the spectral fit. The “relative SNR” of a voxel *i* is defined as 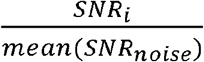, where *SNRnoise* represents the SNR of all non-brain voxels, and is used to access the actual sensitivity enhancement upon incoorporating truncated data.

## Code Availability

The Matlab code to process 3D DMI data, referred to as DMIWizard, is freely available for download from the Yale website or via GitHub (https://github.com/radegraaf/DMIWizard).

## Data Availability

All data produced in the present study are available upon reasonable request to the authors.

## List of abbreviations

CNS –: Central Nervous System
CRLB –: Cramer-Rao Lower Bound
CSF –: CerebroSpinal Fluid
DMI –: Deuterium Metabolic Imaging
DMSO –: dimethylsulfoxide
FID –: Free Induction Decay
FLAIR –: FLuid Attenuated Inversion Recovery
Glc –: Glucose
Glx –: Glutamate+Glutamine
GM –: Gray Matter
GRAPPA –: GeneRalized Autocalibrating Partial Parallel Acquisition
HDO –: deuterium hydrogen monoxide
IDH –: Isocitrate DeHydrogenase
Lac –: Lactate
LW –: Linewidth
MRI –: Magnetic Resonance Imaging
MP-RAGE –: Magnetization Prepared Rapid Gradient Echo
NS –: Number of Slices
PET –: Positron Emission Tomography
SENSE –: SENSitivity Encoding
SNR –: Signal-to-Noise Ratio
SWI –: Susceptibility Weighted Imaging
T_2_W –: T_2_-weighted
TEM –: Transverse ElectroMagnetic
TE –: Echo Time
TR –: Repetition Time
WHO –: World Health Organization
WM –: White Matter

## Acknowledgements

The authors thank Monique Thomas for phantom preparations, and Serena Thaw-Poon for recruiting patients. This research was funded by NIH grant NIBIB R01-EB033764, R01-EB025840, and 2022 ABTA Discovery Grant.

