## Supplementary material for "Parallel detection of multi-contrast MRI and Deuterium Metabolic Imaging (DMI) for time-efficient characterization of neurological diseases": SupplementaryInformation_Medrxiv.docx

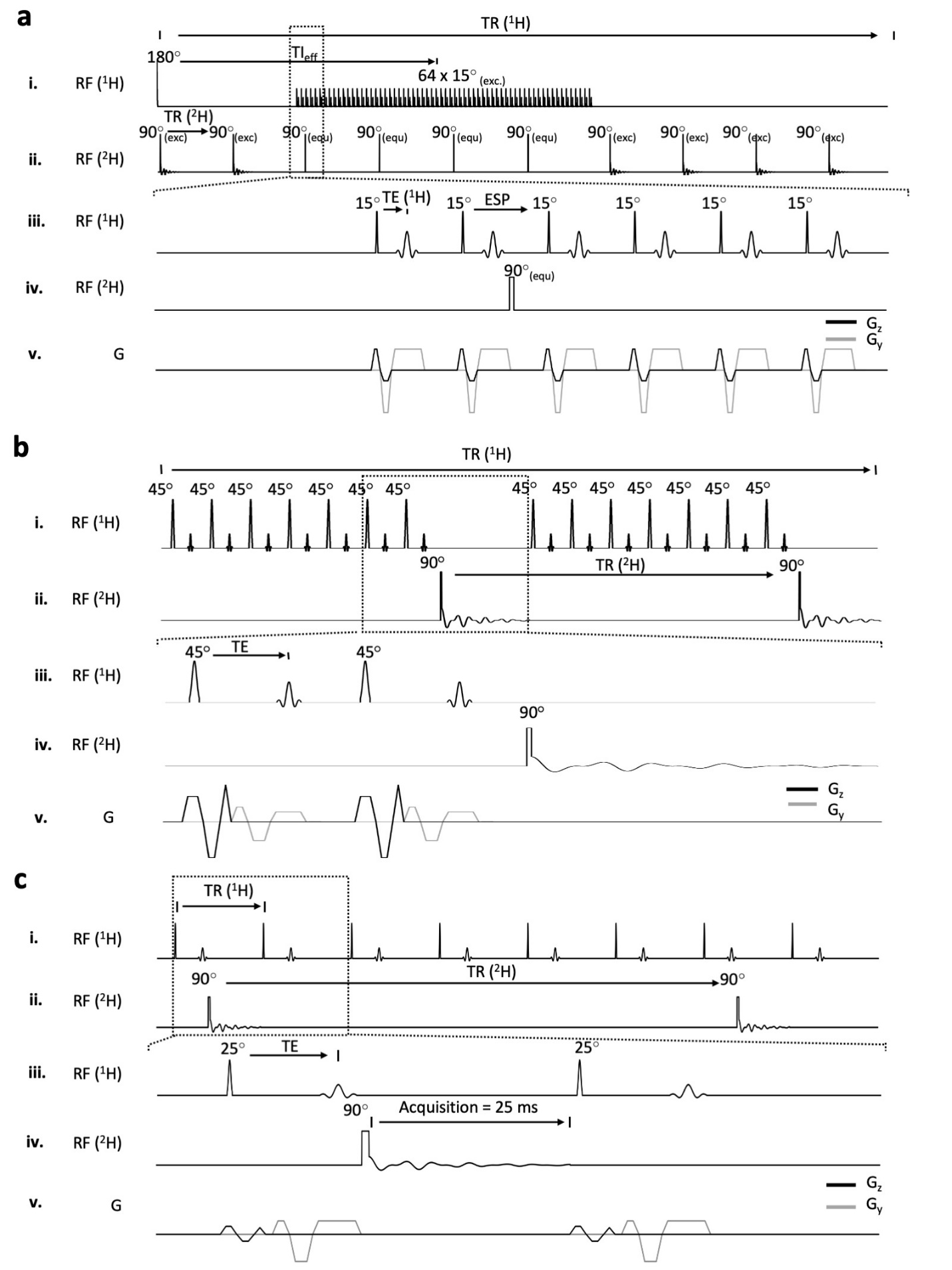

**Figure S1.** **Parallel MRI-DMI sequence design. (a) Parallel MP-RAGE MRI-DMI sequence. (i/iii)** ^1^H pulse acquisition scheme. The tissue section is inverted (TR/TI_eff_= 3140/1240 ms) followed by 64 gradient echoes on z-phase-encoding direction (TE/ESP= 8/20 ms). A total of six ^2^H pulse acquisitions **(ii/iv)** are placed in the dead time during inversion recovery and relaxation with equal TR (= 314 ms). Four ^2^H equilibrium pulses are inserted during ^1^H acquisition to preserve ^2^H steady-state. The gradients used during acquisition are shown in **(v)**. **(b) Interleaved multi-slice SWI MRI-DMI sequence. (i/iii)** ^1^H pulse acquisition scheme. 14 slices are excited (TR/TE = 628/18.5 ms, FA=45^o^). The ^2^H pulse acquisitions **(ii/iv)** are placed every 7 excitations such that DMI TR equals 314 ms. **(v)** Shows the gradients used during acquisition. Flow compensation is applied to all 3 directions. **(c) Interleaved 3D SWI-DMI sequence.** **(i/iii)** ^1^H signal acquisition is achieved using spoiled gradient-echoes (TR/TE= = 52.8/25 ms). **(ii/iv)** ^2^H pulse-acquire elements are placed in the remaining delay following ^1^H signal acquisition, whereby 1250 ^2^H complex points (50 kHz spectral width) were acquired over circa 25 ms. One ^2^H pulse acquisition is inserted for every six ^1^H pulse-acquisitions to achieve a ^2^H TR of 314 ms. **(v)** Magnetic field gradients used during SWI-DMI with flow compensation on the ^1^H slab selection (z) and readout (y) gradients. Phase-encoding gradients in the x and z (MRI) and x, y and z (DMI) directions are not flow-compensated and are not shown. During the total 3D SWI scan time of 10 minutes and 43 seconds, 4 complete DMI datasets can be acquired.

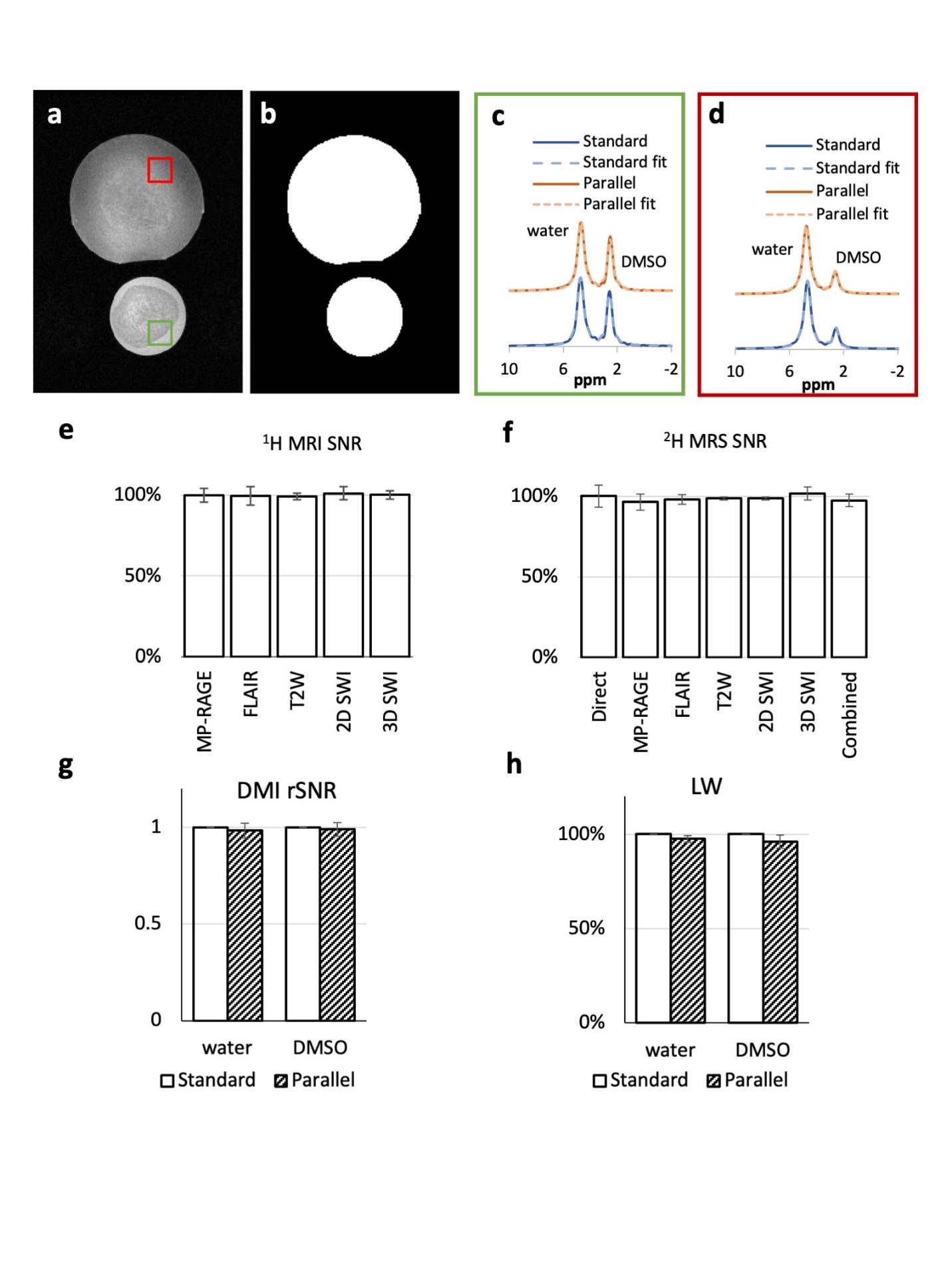

**Figure S2. Parallel MRI-DMI validated in vitro.** Agar phantoms containing D_2_O and DMSO of various concentrations were imaged using the parallel MRI-DMI protocol. The results were compared with the equivalent standard, mono-nuclear acquisitions for validation. **(a)** An example slice of parallel MRI (T_2_W) on phantoms. The contrast in the phantoms is due to inhomogeneous agar distribution. **(b)** The MRI was segmented to generate an MRI image mask. **(c-d)** Localized standard and parallel DMI spectra of two highlighted voxels in **(a)**. **(e)** The SNR of parallel MRIs on the same plane shown in **(a)** were calculated and compared to their equivalent standard control in percentage. The error bars represent the standard deviation of all phantom containing voxels in the image. **(f)** The SNR of ^2^H MRS obtained using five parallel sequences (NA=100) was compared with equivalent ^2^H-only global DMS acquisitions, on individual sequence basis (N=3) and after being combined following the “stitching algorithm” described in the Methods section (N=3). For truncated ^2^H signal, SNR was represented by the standard deviation of the first point of FID. **(g-h).** The rSNR and linewidth (LW) of each localized DMI spectrum was compared voxel-by-voxel in percentage. The error bars in all bar charts represent the standard deviation of all spectra on the demonstrated image plane. One-way or two-way ANOVA was used for statistical analysis. No significant difference (P<= 0.05) was found between the standard and parallel measurements in **(e)**, **(f)**, **(g)**.

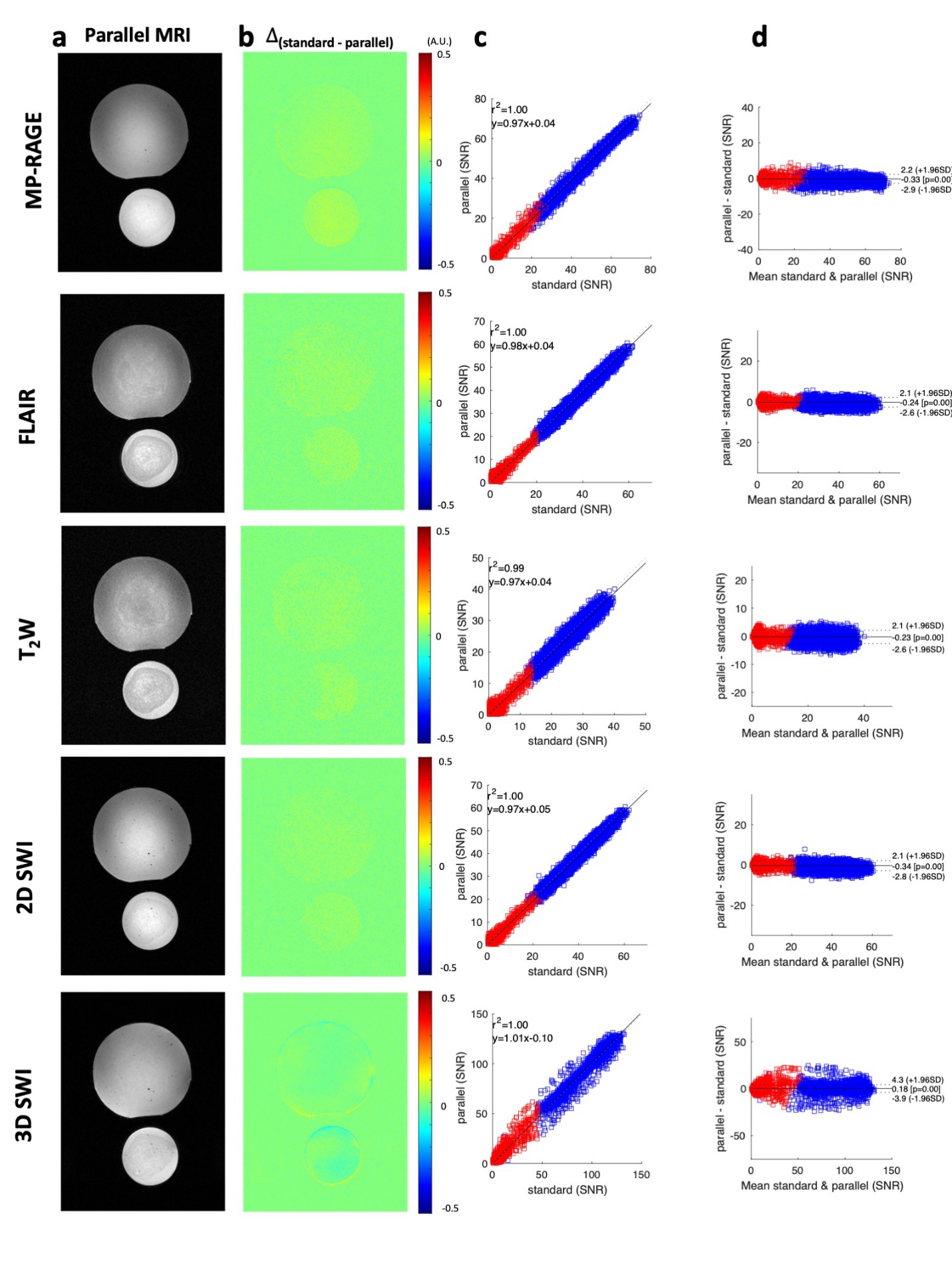

**Figure S3. Comparison of standard and parallel MRI in vitro. (a)** One slice of MRI image on phantoms obtained using the parallel MRI methods (MP-RARAGE, FLAIR, T_2_W, 2D SWI and 3D SWI). **(b)** The SNR difference between standard and parallel MRIs. **(c)** linear regression analysis of parallel vs. standard MRI SNR of the displayed slices on regions with (blue) and without (red) phantom. The fitting results and the goodness of fitting (r^2^) are indicated in the plot. **(d)** Bland-Altman plot of the displayed MRI images. The mean and confidence intervals for 95% limits of agreement (+/- 1.96 S.D.) are indicated in the plot.

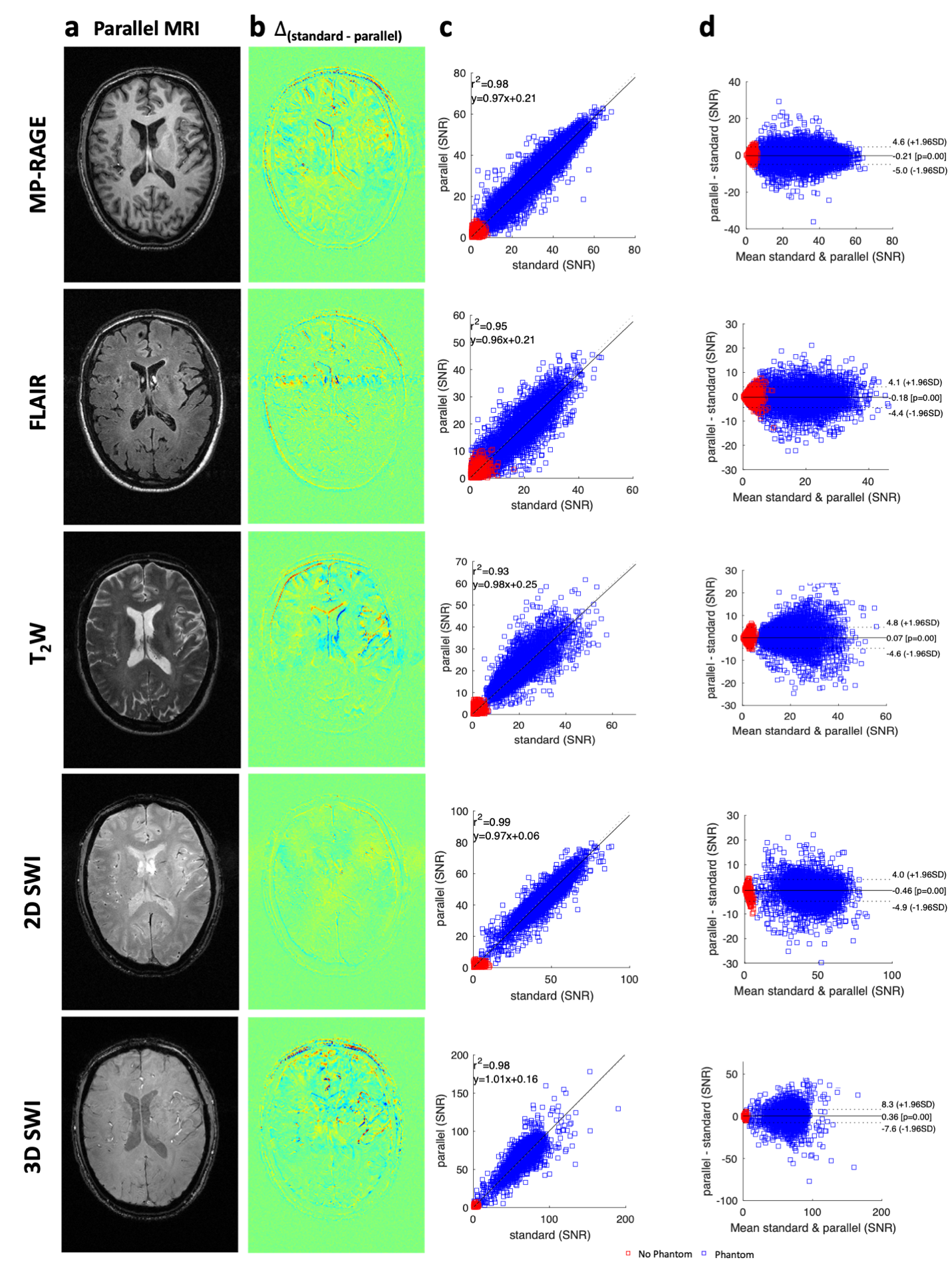

**Figure S4. Comparison of standard and parallel MRI in vivo. (a)** One slice of MRI image in the brain acquired using parallel MRI scans (MP-RARAGE, FLAIR, T_2_W, 2D SWI and 3D SWI). **(b)** The SNR difference between standard and parallel measurements. **(c)** linear regression analysis of parallel vs. standard MRI of the displayed MRI slices on regions with (blue) and without (red) brain tissues. The fitting results and the goodness of fitting (r^2^) are indicated in the plot. **(d)** Bland-Altman plot of the displayed MRI images. The mean and confidence intervals for 95% limits of agreement (+/- 1.96 S.D.) are indicated in the plot.

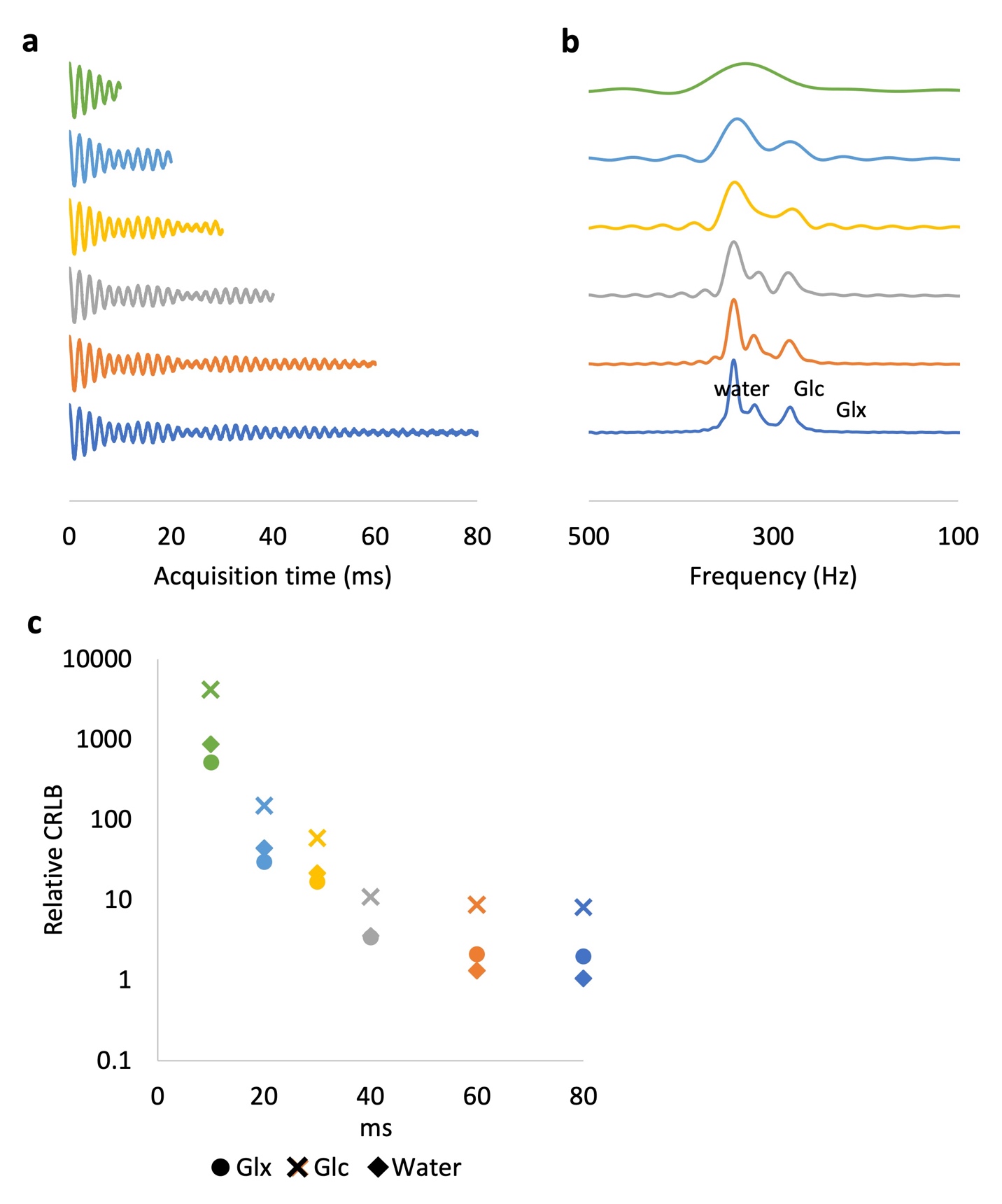

**Figure S5. Effect of FID truncation on quantification accuracy.** **(a)** Simulated ^2^H FID (50 kHz spectral width) with Gaussian noise (𝜎 = 0.05), containing water, glucose (Glc) and glutamate (Glu) calculated over different acquisition periods, from 10 ms (500 points) to 80 ms (4000 points). **(b)**. Spectra corresponding to FIDs in **(a)**. **(c)** The spectra were fitted using linear combination modeling (LCM) with pre-generated basis sets. Cramér-Rao Lower Bounds (CRLB) were calculated, assuming the FIDs at all frequencies have identical phase at time 0. Relative CRLBs were normalized such that CRLB(water)=1 for an infinitely long FID acquisition.

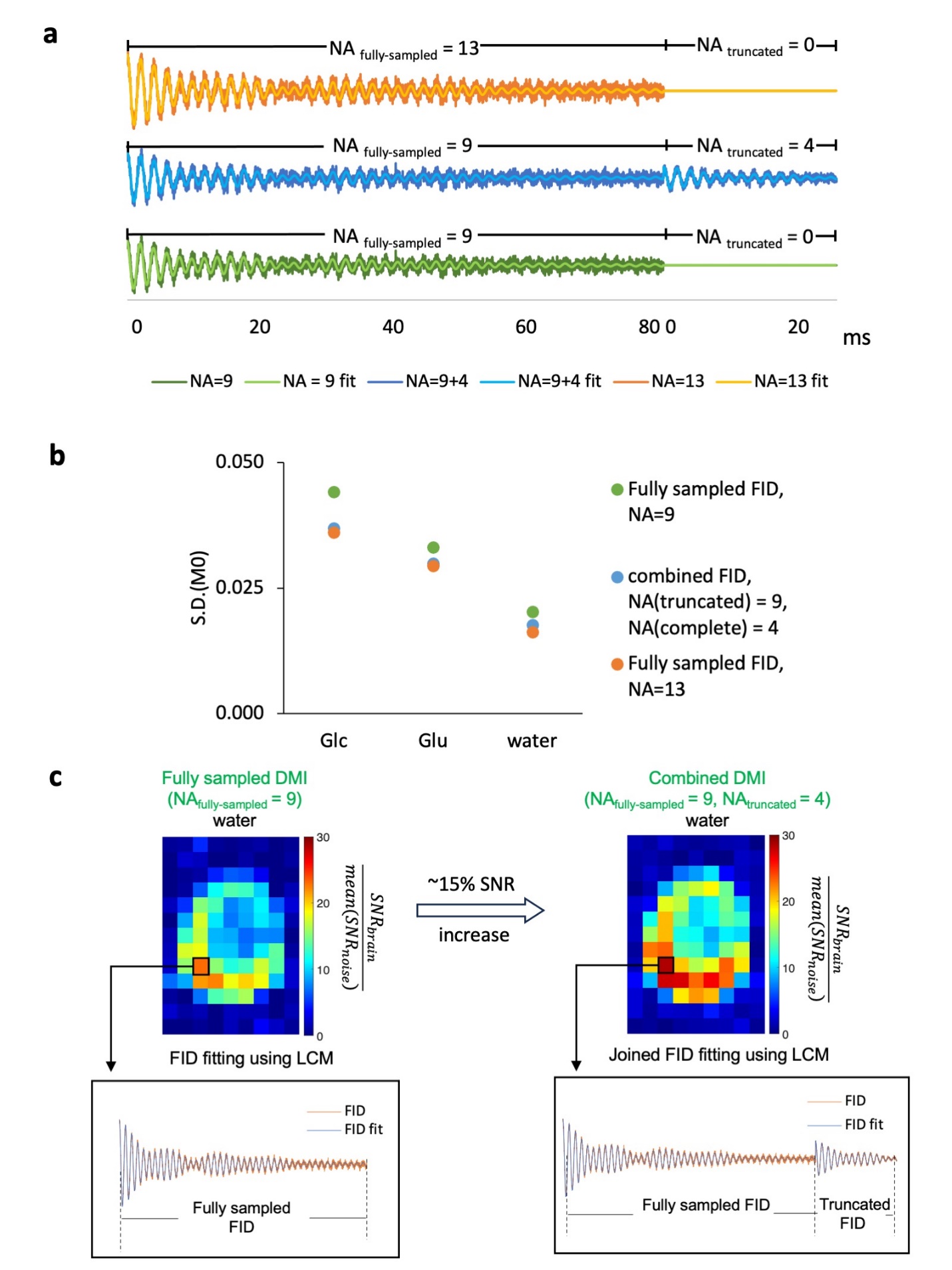

**Figure S6. Sensitivity gain by jointly fitting truncated (NA = 4) and fully-sampled FID (NA = 9). (a)** ^2^H signal (sw = 50KHz) containing water, glucose (Glc) and glutamine+glutamate (Glx) peak, with linewidth resembling in vivo measurements (LW_water_=10Hz, LW_Glc_=15 Hz, LW_Glx_=15Hz), are simulated with Gaussian distributed noise (𝜎 = 0.4) to generate fully sampled (80 ms, np=4000, NA =9, 13) and truncated FIDs (25 ms, np=1250, NA = 4). For joined fitting, full length and truncated signal were combined and LCM was performed using weighted nonlinear least-square fitting, with weighting of $1/\sqrt{N_{i}}$**. (b)** Monte-Carlo simulation (N _simulations_ =100) was performed to evaluate and compare the fitting accuracy of full-length signal and the combined signal. The standard deviation (S.D.) of fitted M0 indicates that incorporating truncated signal with fully-sample data will significantly improve the fitting, hence increase DMI sensitivity. **(c)** n vivo DMI were calculated with and without addition of truncated FID, followed by calculation of rSNR ( = $\frac{{SNR}_{brain}}{mean({SNR}_{noise})}$) on the water peak to show that incorporation of truncated data improves the sensitivity of DMI signal.

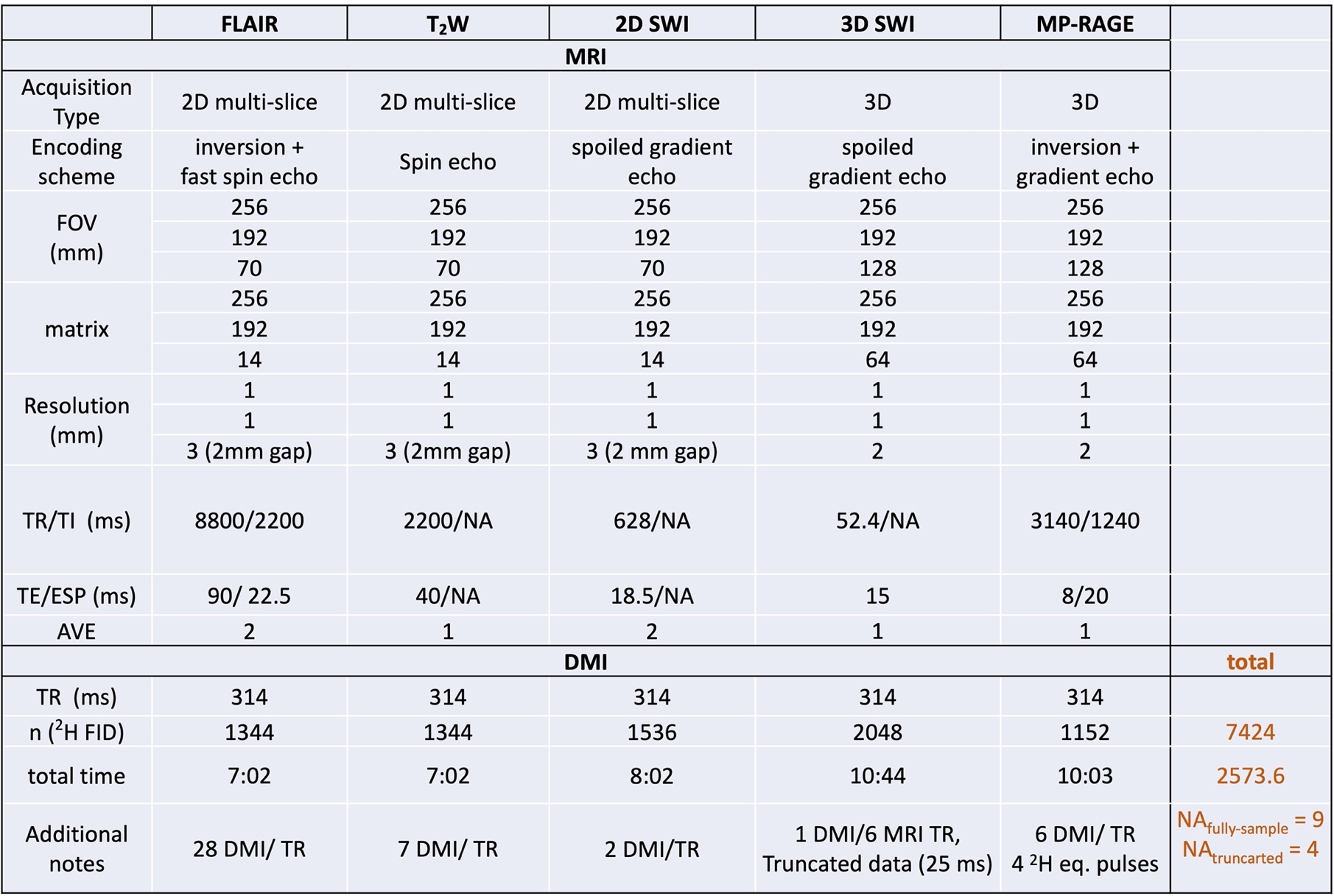

**Table S1.** Detailed information and acquisition parameters of the parallel MRI-DMI sequences.
